# Identification and development of Tetra-ARMS PCR-based screening test for a genetic variant of OLA1 (Tyr254Cys) in the human failing heart

**DOI:** 10.1101/2023.10.16.23296746

**Authors:** Praveen K Dubey, Shubham Dubey, Sarojini Singh, Purnima Devaki Bhat, Steven Pogwizd, Prasanna Krishnamurthy

## Abstract

Obg-like ATPase 1 (OLA1) protein has GTP and ATP hydrolyzing activities and is important for cellular growth and survival. The human OLA1 gene maps on chromosome 2, at the locus 1q31, close to the Titin (TTN) gene, which is associated with familial dilated cardiomyopathy (DCM). In this study, we found that expression of OLA1 was significantly downregulated in human failing heart tissue (HF) as compared to in non-failing heart tissues (NF). Moreover, using the Sanger sequencing method, we characterized the human OLA1 gene and screened genetic mutations in patients with heart-failing and non-failing. Among failing and non-failing heart patients, we found a total of 15 mutations, including two transversions, one substitution, one indel, and eleven transition mutations in the *OLA1* gene. All the mutations were intronic except for a non-synonymous mutation, 5144A>G, resulting in 254Tyr>Cys in exon 8 of the *OLA1* gene. Furthermore, haplotype analysis of these mutations revealed that these single nucleotide polymorphisms (SNPs) are linked to each other, resulting in disease-specific haplotypes. Additionally, to screen for the 254Tyr>Cys point mutation, we developed a cost-effective, rapid genetic screening PCR test that can differentiate between homozygous (AA and GG) and heterozygous (A/G) genotypes. Our results show that this test can be used as a genetic screening tool for human cardiomyopathy. These findings have important implications for the diagnosis and treatment of cardiomyopathy.

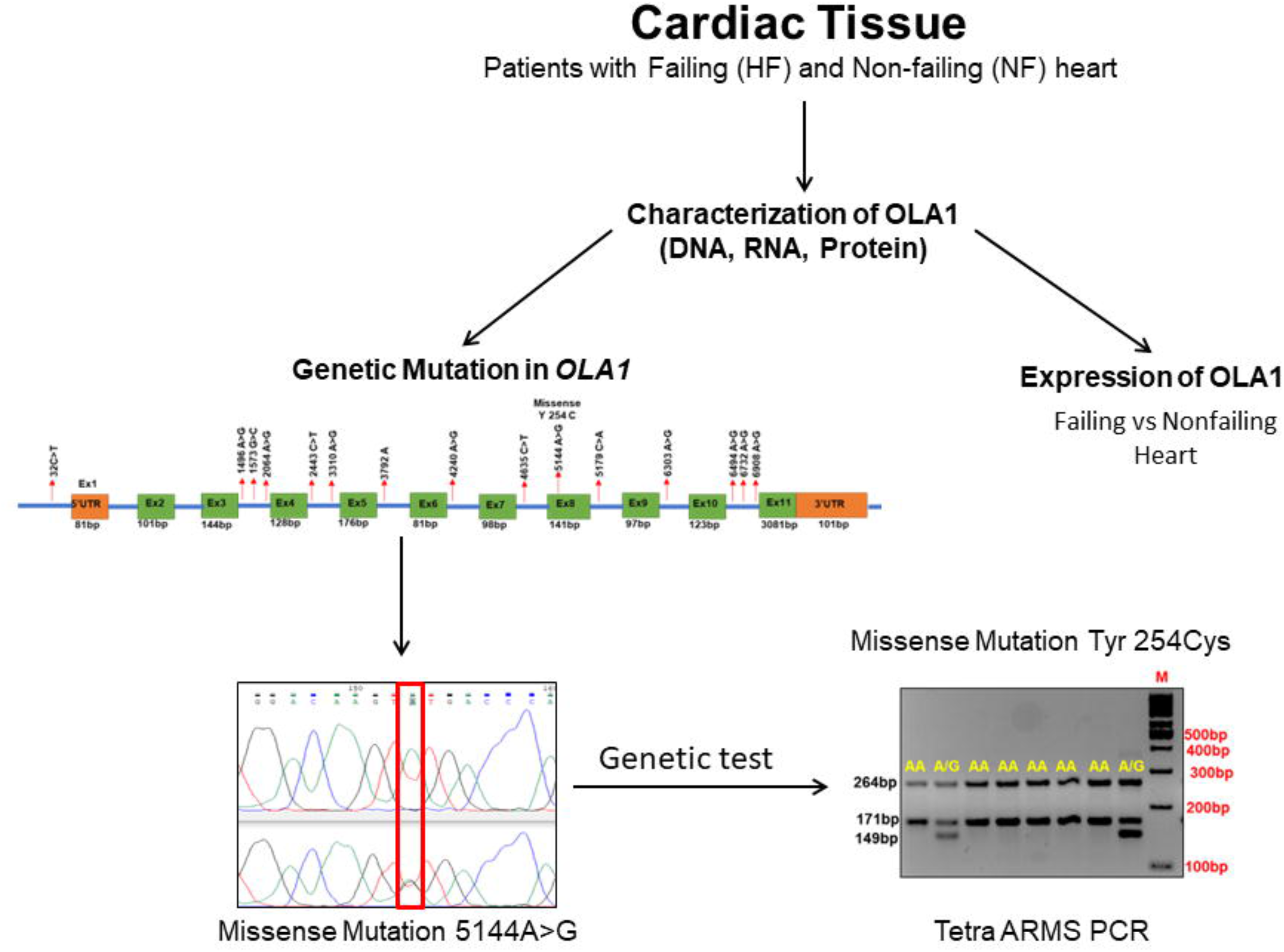

## Introduction

Obg-like ATPase 1 (OLA1) is a member of the GTPase family of proteins, exhibiting both GTPase and ATPase activities. It is highly expressed in cancer cells and is associated with poor survival [1,2]. In cancer cells, increased expression of OLA1 inhibits apoptosis by interacting with breast cancer-associated gene 1 (BRCA1) and BRCA1-associated RING domain protein (BARD1) [3–5]. OLA1 also interacts with eIF2α to form a ternary complex for regulating protein translation and cell proliferation [6]. A recent study in mice showed that OLA1 interacts with Hsp70 to stabilize mitochondrial superoxide dismutase 2 (SOD2) in pulmonary smooth muscle cells; a loss of function for OLA1 caused SOD2 deficiency, resulting in the increased expression of the X-linked inhibitor of apoptosis (*XIAP*) gene, and increased proliferation of pulmonary SMCs [5,7].

Proteomic and functional studies in human hypertrophic cardiomyopathy (HCM) patients show that expression of several genes in cardiac tissue including *OLA1* was downregulated [8]. Our previous study also found that treating mice with angiotensin II (AngII) altered the expression of OLA1, resulting in the phosphorylation of GSK3β/β-catenin pathway involved in the development of cardiac hypertrophy [4]. Furthermore, studies have shown that deletion of *OLA1* in mice leads to a significant increase in heart weight compared to body weight [6], indicating it’s potential impact on cardiac structure and function.

Although mutations in more than 40 genes are known to cause DCM in humans, the role of *OLA1* mutations in human heart failure has not been studied yet. In this study, we aimed to screen genetic mutations in the *OLA1* gene and investigate their association with heart failure in human patients. Additionally, we developed a cost-effective and rapid PCR-based genetic screening test. Based on our findings reported in this study, future investigations will enhance our understanding of the pathogenesis of cardiomyopathy and may provide novel therapeutic targets for the treatment of heart failure.

## Materials and methods

### Patient samples

We obtained anonymized human tissue samples from non-failing heart (n=5; NF1-5) and failing heart (n=5; HF6-10) from tissue repository at the University of Alabama at Birmingham (UAB). Patient identity is not known to the research group. The study protocols were approved by the UAB Institutional Review Board.

### Mouse heart tissue samples

Mice tissue samples were collected from Wild-type C57BL/6J mice (10-12 weeks old) that were purchased from Jackson Laboratory, Bar Harbor, ME. All protocols were approved by the Institutional Animal Care and Use Committee (IACUC) of the University of Alabama at Birmingham.

### RNA isolation and cDNA synthesis

Total RNA was isolated from ~50 mg of human failing and non-failing heart tissues using the TRIzol method (Invitrogen) and purified with the Qiagen RNA extraction kit (cat# 74106, Qiagen) according to our published protocol [9–11]. RNA quality and quantity were assessed with Nanodrop, and 1 µg of RNA from each sample was reverse transcribed using the RevertAid First Strand cDNA Synthesis Kit (cat# K1691, ThermoFisher Scientific). Specific primers (Supplementary Table 1) were used for performing qPCR analyses on a QuantStudio 3 system (Applied Biosystems, ThermoFisher Scientific) using the PowerUp™ SYBR™ Green Master Mix (cat# A25778, ThermoFisher Scientific). Target gene expression was normalized to housekeeping genes (18S rRNA or β-actin or GAPDH) and presented as fold change versus control.

### DNA isolation

Total DNA from human failing and non-failing heart tissues (~5 mg) was isolated using the Qiagen DNAeasy kit (cat# 69504, Qiagen) according to the manufacturer’s instructions. The quality and quantity of the isolated DNA were measured using Nanodrop.

### Primers designing and PCR amplification

To screen polymorphisms in the *OLA1* gene, nine sets of primers (Supplementary table 1) were designed using online Primer Blast server of NCBI based on available human *OLA1* gene sequences from the ensemble genome browser (Transcript: ENST00000284719.8). Different fragments of PCR products covering all exons along with partial introns were amplified from isolated genomic DNA from both failing and non-failing patient heart tissue. Before sequencing, all PCR products were optimized for amplification and sequencing performance to ensure reliable screening for mutations in the *OLA1* gene. Details of the PCR primers sequence, along with their annealing temperature and size, are provided in Supplementary Table 1. For each PCR reaction, a total volume of 20 μl containing 20-50ng genomic DNA, 10 pmole of each primer, along with 2X PCR Master Mix (Cat.no. PRM7505, Promega) was used. After an initial denaturation step (95°C for 3 min), samples were subjected to 32 cycles of PCR consisting of 94°C for 30s, primer-specific annealing temperature (Supplementary Table 1) for 30 sec, and 72°C for 90 sec, followed by a final extension for 7 min at 72°C. Amplified products were then checked on a 1.5% agarose gel, and images were captured using Bio-Rad Gel doc with a UV transilluminator.

### DNA Sequencing and Analyses

Prior to Sanger sequencing, the amplified PCR products were enzymatically cleaned up using ExoSAP-IT™ PCR Product Cleanup Reagent (Cat no. 78200. 200.UL, Thermofisher). Subsequently, the cleaned PCR products were sent for Sanger sequencing at the UAB core facility. Each PCR product was sequenced with its respective PCR primer using the fluorescent dideoxy terminator method of cycle sequencing (ABI Prism BigDye Terminator v3.1 Cycle Sequencing kits using 1/16 of the “standard” protocol). The sequencing reactions were purified using the Applied Biosystems X Terminator purification system as per the manufacturer’s protocols. After purification, sequencing reactions were run on a 3730xl DNA Analyzer (Applied Biosystems Division, Foster City, CA) following Applied Biosystems’ protocols. The sequence of each DNA sample was manually checked using the Chromas Lite program (http://www.technelysium.com.au/chromas_lite.html) and further subjected to multiple alignments to identify nucleotide variations, using the MegAlign program of the Lasergene software. Finally, different fragments of the *OLA1* gene sequences were assembled using the SeqMan program of Lasergene and finally submitted to NCBI GenBank

### Genetic screening test for the *OLA1* point mutation (254 Tyr>Cys)

For genotyping of the non-synonymous SNP A5144G, which results in the amino acid change 254Tyr>Cys in Exon8 of the *OLA1* gene, a tetra-primers ARMS-PCR-based screening protocol was developed using four different sets of primers (shown in supplementary Table 1) following previously described protocols [12–14]. The primers were designed from the human *OLA1* sequence (accession no. ON073791.1) using web-based software available at http://cedar.genetics.soton.ac.uk/public_html/primer1.html and synthesized through IDT. Nucleotide-specific amplification of the genotype protocol was standardized and validated by Sanger sequencing. PCR was performed on isolated genomic DNA samples in a 20 µl reaction volume containing approximately 25-50 ng of template DNA, 0.5 µl of 10 pmol of each inner primer, 0.1 µl of 10 pmol of each outer primer, and 2X PCR Master Mix (Cat.no. PRM7505, Promega). Amplification was carried out using the following PCR conditions: 95°C for 3 min, followed by 32 cycles of 94°C for 30 sec, annealing at 58°C for 30 sec, and 72°C for 1 min, followed by a final extension at 72°C for 5 min. Samples were checked on ethidium bromide-stained 2% ultra-pure agarose gel, and genotypes were recorded.

### Protein extraction and Western blot analysis

Protein lysates from human failing and non-failing hearts and different tissues from mice were prepared using the RIPA Cell Lysis Buffer (cat# J63324, Alfa Aesar) supplemented with Halt^TM^ protease inhibitor (cat# 87786, ThermoFisher Scientific) cocktails and phosphatase inhibitor (cat# P5726, Millipore Sigma) following our lab published protocol. Equal amounts of proteins were loaded on AnyKD Mini-PROTEAN TGX stain-free protein gels (cat# 4569033, Bio-Rad Laboratories) and transferred to polyvinylidene difluoride (PVDF) membranes using Trans-Blot Turbo transfer system (Bio-Rad Laboratories). The immunoblots were quickly washed with water and blocked with 5% non-fat milk for an hour. After a quick wash with water, the blots were incubated with primary antibodies against OLA1 (cat# 16371-1-AP, Proteintech) and β-Tubulin (cat# 10094-1-AP, Proteintech). After washing, blots were incubated with secondary HRP-conjugated antibodies against mouse (cat# SA00001-1, Proteintech) and rabbit (cat# SA00001-2, Proteintech) at room temperature for 1 hour on a shaker. Images were acquired using a ChemiDoc™ Touch Imaging System (Bio-Rad, USA) using an enhanced chemiluminescence (Pierce) detection system.

### Cell culture and protein fractionation

Human ventricular cardiomyocyte cell line (AC16; Cat# SCC109) were purchased from Millipore Sigma and cultured in DMEM/F12 (Cat# D6434, Sigma-Aldrich) supplemented with 2 mm L-Glutamine (Cat# TMS-002-C EMD Millipore), 12.5% fetal bovine serum (FBS; Cat# ES-009-B, EMD Millipore) and 1% Penicillin-Streptomycin Solution (Cat# 15140122, Thermo Fisher Scientific) in a humidified incubator at 37°C with 5% CO2. Cells were seeded in 10 cm dish for cytoplasmic and nuclear fractionation. NE-PER™ Nuclear and Cytoplasmic Extraction kit (cat# 78835, Thermofisher Scientific) was used for cytoplasmic and nuclear fractionation. Protease and phosphatase inhibitors were added before use. Fractionated proteins were separated on an SDS-PAGE, the proteins were transferred to membrane and Western analysis was done as above with antibodies against OLA1, GAPDH (cytoplasmic marker) and Lamin B1 (Nuclear marker).

### Cohort Study

To confirm our finding of the genetic mutation in *OLA1*, we cross-referenced it with a large dataset, such as the REGARDS study. This National Institutes of Health (NIH)-funded cohort study was conducted between 2003 and 2007 with 30,239 participants involving representations from various demographic backgrounds across the continental United States. We also utilized genetic variant data set available on Ensembl (https://useast.ensembl.org/Homo_sapiens/Transcript/ProtVariations?db=core;g=ENSG00000138430;r=2:174072447-174248599;t=ENST00000284719).

## Results

### Expression of OLA1 was downregulated in human failing heart

OLA1 has been shown to play a pivotal role in the progression of cancer [6] and heart disease [2,4,8]. In this study we assessed the expression of OLA1 (mRNA and protein) in human failing (HF) and non-failing (NF) heart. Interestingly, our qPCR analysis indicated a significant downregulation of OLA1 mRNA expression in failing heart tissues compared to non-failing heart tissue (**Figure 1A**). Furthermore, Western analysis of isolated proteins from the same tissues revealed a concurrent reduction in OLA1 protein expression in failing hearts relative to non-failing hearts (**Figure 1B**). Densitometric analysis, normalized with the respective housekeeping gene (β-Tubulin), confirmed a substantial decrease in OLA1 protein levels in failing hearts (**Figure 1C**). These findings collectively suggest that both mRNA and protein levels of OLA1 were significantly downregulated in failing hearts compared to non-failing hearts (**Figure 1A, B & C**).

**Figure 1.**
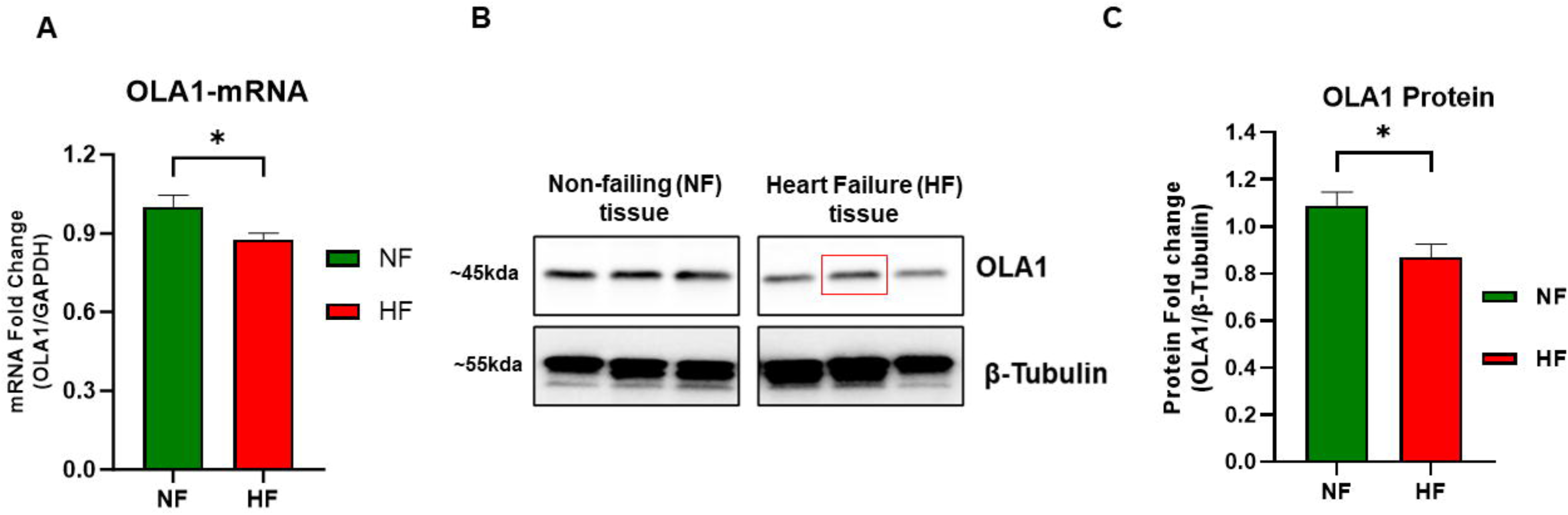
Expression of OLA1 in human heart failing (HF) and non-failing heart (NF). (A) mRNA quantification of the *OLA1* gene using quantitative real-time PCR in a subset of the Failing heart tissue (n=5; HF) and non-failing heart tissue from control subjects (n=5; NF). Glyceraldehyde 3-phosphate dehydrogenase (GAPDH) served as internal controls for normalization. Data shown in relative fold change. (B) Representative immunoblot depicting OLA1 protein expression in proteins extracted from human hearts displaying severe left ventricular dysfunction (non-ischemic, heart failing, n=5, HF) and non-failing control subjects (n=5). The red square denotes a sample obtained from heart carrying an OLA1 variant (254Tyr>Cys). (C) The densitometric quantification of immunoblots. The data are presented as fold change and expressed as mean ± SEM. Data were analyzed using a two-tailed unpaired student *t*-test, with significance set at *p < .05.

To determine the tissue-specific expression pattern of the *OLA1* gene and the subcellular localization of the OLA1 protein, we examined OLA1 expression across various mouse tissues via immunoblotting, including Brain, Adipose Tissue, Lung, Kidney, Spleen, Liver, Skeletal Muscle, and Heart. The results revealed a ubiquitous expression of OLA1 protein in all tissues with a slightly higher expression in Skeletal Muscle and Heart (**Supplementary Figure S1A, B**). Furthermore, subcellular fractionation of cytoplasmic and nuclear proteins from immortalized human ventricular cardiomyocytes (Ac16 cells) demonstrated that OLA1 primarily localizes in the cytoplasmic fraction compared to lower levels in the nucleus (**Supplementary Figure S1C**). These data suggests that OLA1 might be primarily exerting its function within the cytosol of cells, perhaps more so in energy-demanding tissues such as skeletal muscle and heart.

### Characterization of the human *OLA1* gene

To comprehensively characterize the human *OLA1* gene at both the mRNA and genomic levels, we amplified both mRNA and genomic sequence of the *OLA1* gene using RNA and DNA extracted from human heart tissue. For mRNA characterization, we carried out the amplification of the entire Open Reading Frame (ORF) of the *OLA1* mRNA and subsequently cloned it into the pCDNA3.1 TA cloning vector. Using Sanger sequencing various fragments of the *OLA1* sequence were assembled and subjected to NCBI Blast analysis to assess their homology. Upon confirmation of the sequence accuracy for the human *OLA1* gene, the *OLA1* mRNA was translated into amino acids and deposited to the NCBI database under the accession number ON073790.1.

Our sequence analyses unveiled that the coding DNA sequence (CDS) of the human *OLA1* gene spans 1191 base pairs, ultimately encoding a 396 amino acid protein, in agreement with findings in other species (**Figure 2A**). Structural analysis using PsiPred for 2D and 3D structure of the human OLA1 translated protein showed the presence of 13 strands, 16 helices, and 23 coils (**Figure 2B, C**).

**Figure 2.**
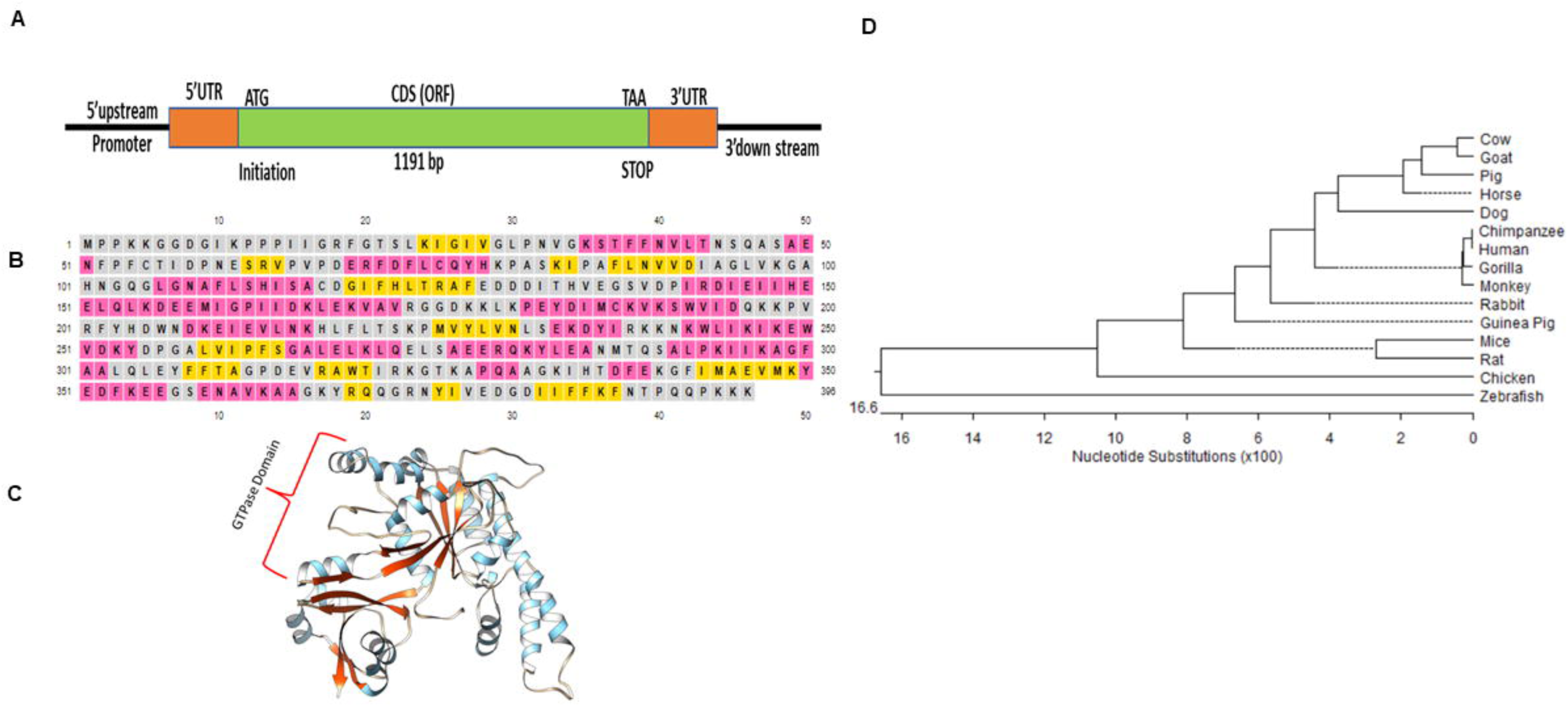
The characterization of Human *OLA1* Gene. Panel (A) shows the amplified mRNA of the *OLA1* gene, highlighting the coding sequence (CDS) in green (1191bp), along with the 5’-UTR and 3’-UTR. Panel (B) depicts the translated amino acids of the *OLA1* gene, with the secondary structure indicated by helices (pink), coils (brown), and strands (yellow). Panel (C) illustrates the tertiary structure of the OLA1 protein. Panel (D) presents the phylogenetic relationship of the human OLA1 gene with other species at the nucleotide level.

Furthermore, phylogenetic analysis based on both nucleotide and amino acid sequences, indicated that the human *OLA1* gene shares a close evolutionary relationship with chimpanzees and monkeys, with the lowest similarity observed in zebrafish (89%) (**Figure 2D**). Percentage homology among different species and phylogeny shown in **Supplementary figure S2 A, B, C**. These findings provide valuable insights into the gene characteristics and evolutionary connections of the human *OLA1* gene among various species.

### Genomic organization of the human *OLA1* gene

For genomic characterization of the human *OLA1* gene, different fragments of the *OLA1* genomic sequence were amplified using different set of primers (Supplementary Table 1) and confirmed based on their respective size with agarose gel analysis (Supplementary Figure 3A). Different fragments of the *OLA1* gene were sequenced and assembled. The Open reading frame (ORF) of *OLA1* was determined by aligning with the human *OLA1* mRNA sequence and further validated using the online NCBI Splign program. This analysis revealed that the *OLA1* gene is composed of 11 exons, with the start codon located in exon 2 and the stop codon in exon 11 (Supplementary Figure 3B). The length of each exon is shown in the Supplementary Figure 3B and has been submitted to the NCBI gene data bank with an accession number ON073791.1. Furthermore, we also showed that the intronic sequence of the *OLA1* gene is relatively long, spanning over 100,000 base pairs. This information is of potential importance for determining the regulatory mechanisms that control the expression of the *OLA1* gene.

**Figure 3.**
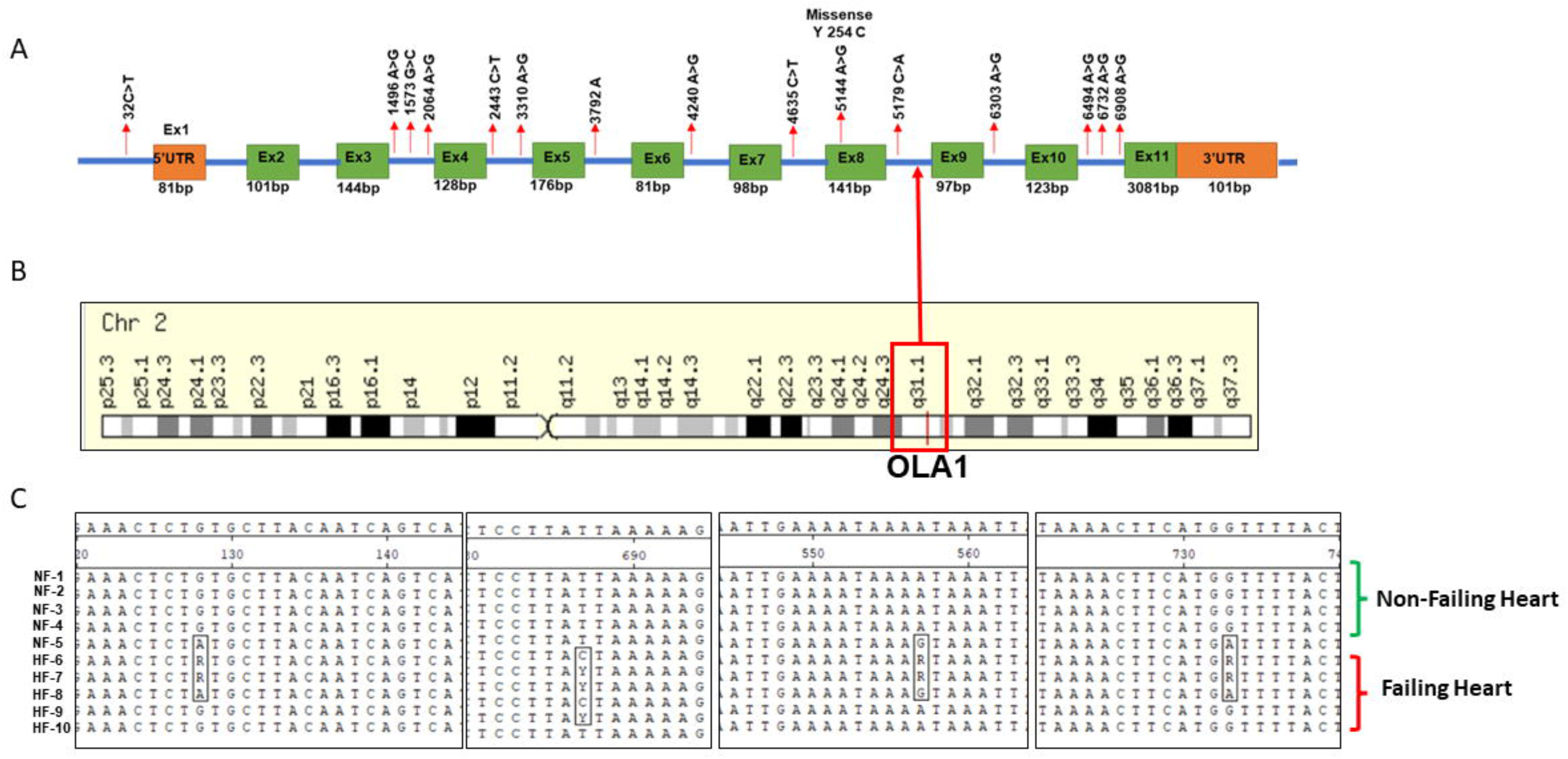
Mutations in the *OLA1* gene detected in both failing and non-failing patients. Panel (A) illustrates the distribution of mutations within exons and introns. Panel (B) shows the location of the *OLA1* gene on human chromosome 2, Locus 2q31.1, which is associated with DCM in humans, as depicted in NCBI MapViewer. Panel (C) displays a MegaAlignment of nucleotide sequences, demonstrating that the *OLA1* mutations are interconnected and exhibit distinct patterns between failing (HF 6-10) and non-failing heart (NF1-6) patients.

### Expression of the *OLA1* splice variant in Human heart tissue

OLA1 has been shown to play a critical role in the progression of cancer and heart diseases [4,6,8,17]. Interestingly, several splice variants of the *OLA1* gene have been reported in ensemble human genome browser and the NCBI bank. To identify splice variants in the human heart, we designed primers targeting each of the variants, after aligning with the ensemble human data bank. To find the splice variants, we amplified PCR product from reverse transcribed RNA isolated from human heart samples (Supplementary Figure S4). Our analysis revealed four distinct splice variants (202, 203, 205, and 209; Supplementary Figure S4) expressed in the human heart. Future studies need to explore specific roles for these splice variants in cancer and heart disease.

**Figure 4.**
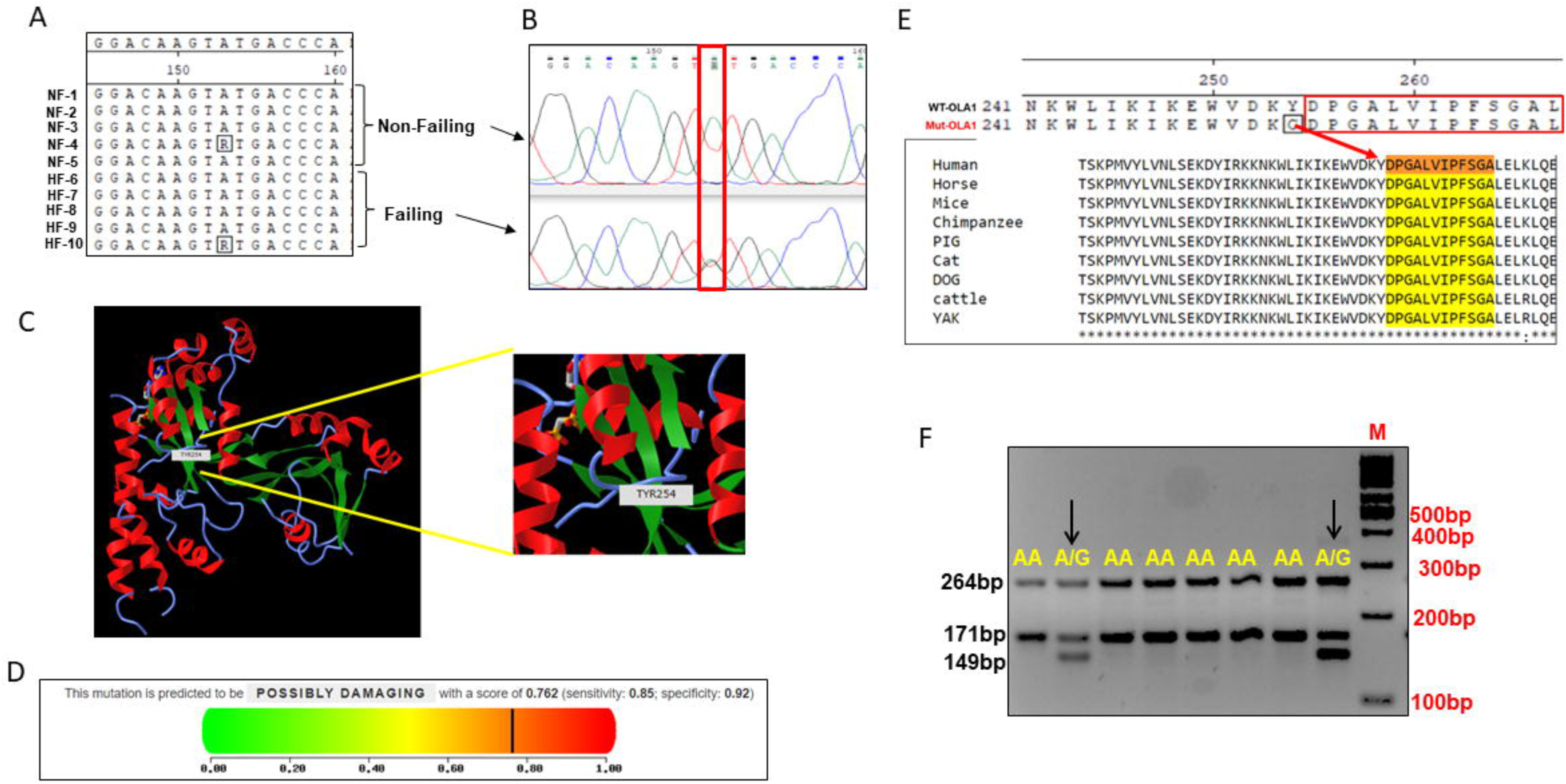
Non-synonymous Mutation (254Tyr>Cys) in Exon 8 of the OLA1 Gene. Panel (A) provides a MegaAlignment, demonstrating a heterozygous (R, A>G) variation in exon 8 of the *OLA1* gene. Panel (B) displays a chromatogram showing the shift from a homozygous AA to a heterozygous A>G peak. Panel (C) presents the Phyphen-2 analysis of the 254 Tyr>Cys mutation, indicating its detrimental impact on protein function. Finally, panel (D) shows the three-dimensional structure of the OLA1 mutation. (E) Depicts the conservation of the mutation in OLA1 (254Tyr>Cys) across different species. (F) Demonstrates the development of a PCR-based assay for screening the *OLA1* mutation (5144A>G).

### Detection of mutations in the *OLA1* gene

In most cases (70-80%), familial Dilated Cardiomyopathy (DCM) is inherited as an autosomal dominant disease, wherein a single copy of the altered gene is sufficient to precipitate the disorder. Previous studies have demonstrated a strong association between a locus on chromosome 2q31, designated as CAD1G, and familial DCM in humans. Notably, this locus encompasses the pivotal cytoskeletal muscle protein TITIN [18]. Genetic mutations within the TITIN gene lead to DCM by compromising the contractile function critical for myofibrillar elasticity and structural integrity [19]. Intriguingly, the *OLA1* gene mapped to the same locus on chromosome 2q31, in close proximity to the *Titin* gene. Alterations in the expression of *OLA1* have been identified in samples from human heart failure cases, as well as in cardiac tissue of mice following ANGII administration [4,8].

Studies in human cancer have showed that mutations in the *OLA1* gene correlate with swift disease progression and unfavorable prognosis. A recent investigation in a breast cancer cell line demonstrated a point mutation (E168Q) in the *OLA1* gene, which impaired its ability to bind BRCA1 for the regulation of centromere amplification [3]. Additionally, research involving atherosclerosis patients has revealed several mutations in the *OLA1* gene, with five of them exhibiting a strong association with atherosclerosis development [17].

To investigate the presence of genetic mutations in the *OLA1* gene associated with heart disease, we performed amplification of the *OLA1* gene, encompassing all exons as well as portions of the adjacent introns, from DNA isolated from human failing (n=5) and non-failing (n=5) heart tissues. Through Sanger sequencing and subsequent sequence analysis, we identified a total of 15 mutations, comprising two transversions, one substitution, one deletion, and 11 transition mutations in the OLA1 gene (**Figure 3**, **Table 1**). The distribution of these Single Nucleotide Polymorphisms (SNPs) among failing and non-failing heart patients is shown in **Supplementary Figure S5**. Remarkably, all the identified mutations were found to be intronic, except for one non-synonymous mutation 5144A>G, resulting in 254Tyr>Cys, located in exon 8 of the *OLA1* gene. Furthermore, we conducted an in-depth analysis of the genotype and allelic frequencies of these identified SNPs in the human *OLA1* gene, as outlined in **Table 1**. The allele frequencies ranged from 0.1 to 0.35 among both failing and non-failing heart patients.

**Table 1.**
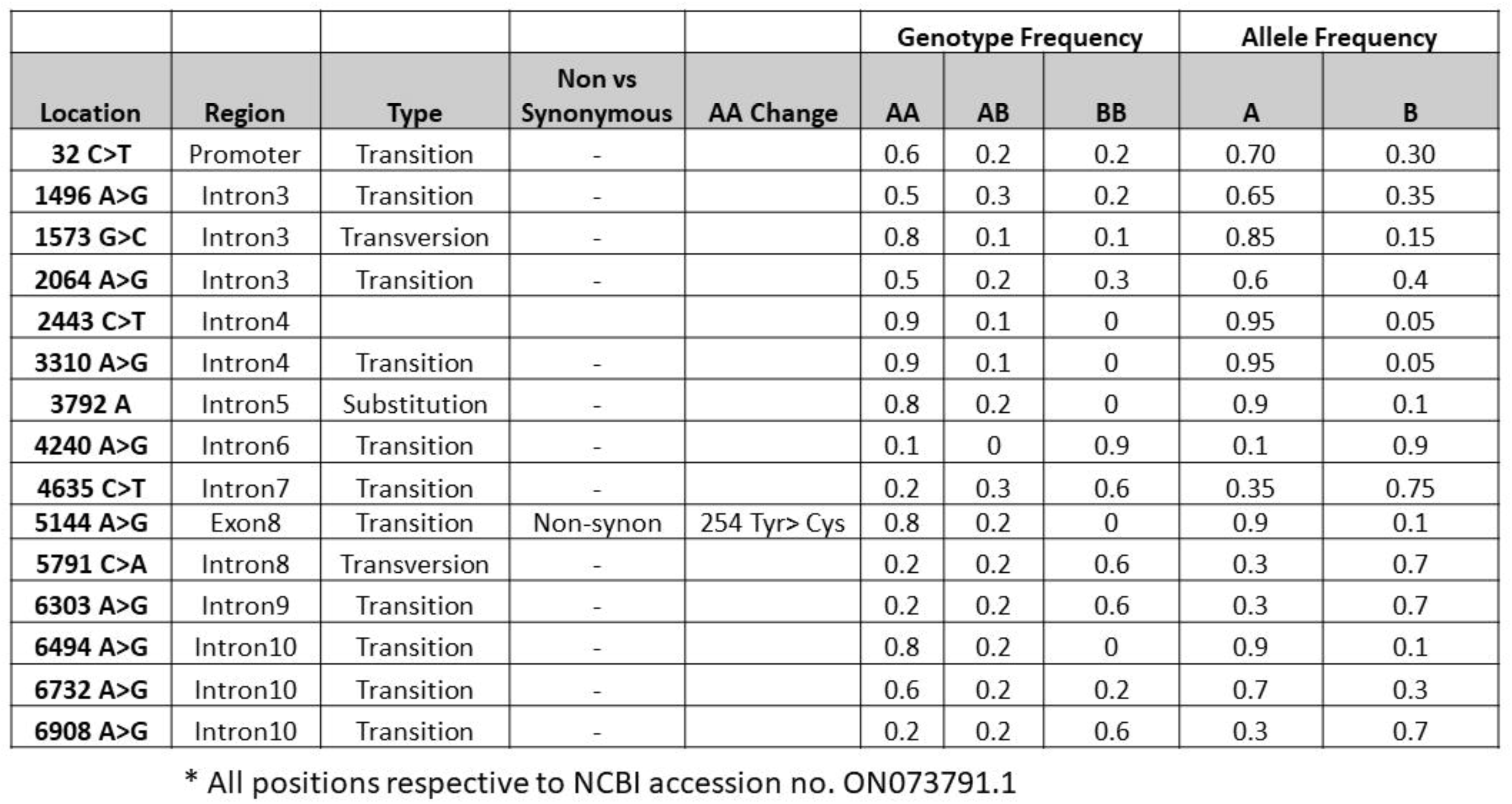
Mutations in the OLA1 gene and its allelic and genotype frequency in human heart from failing and non-failing patients.

|  |  |  |  |  | Genotype Frequency |  |  | Allele Frequency |  |
| --- | --- | --- | --- | --- | --- | --- | --- | --- | --- |
| Location | Region | Type | Non vs<br>Synonymous | AA Change | AA | AB | BB | A | B |
| 32 C>T | Promoter | Transition | - |  | 0.6 | 0.2 | 0.2 | 0.70 | 0.30 |
| 1496 A>G | Intron3 | Transition | - |  | 0.5 | 0.3 | 0.2 | 0.65 | 0.35 |
| 1573 G>C | Intron3 | Transversion | - |  | 0.8 | 0.1 | 0.1 | 0.85 | 0.15 |
| 2064 A>G | Intron3 | Transition | - |  | 0.5 | 0.2 | 0.3 | 0.6 | 0.4 |
| 2443 C>T | Intron4 |  |  |  | 0.9 | 0.1 | 0 | 0.95 | 0.05 |
| 3310 A>G | Intron4 | Transition | - |  | 0.9 | 0.1 | 0 | 0.95 | 0.05 |
| 3792 A | Intron5 | Substitution | - |  | 0.8 | 0.2 | 0 | 0.9 | 0.1 |
| 4240 A>G | Intron6 | Transition | - |  | 0.1 | 0 | 0.9 | 0.1 | 0.9 |
| 4635 C>T | Intron7 | Transition | - |  | 0.2 | 0.3 | 0.6 | 0.35 | 0.75 |
| 5144 A>G | Exon8 | Transition | Non-synon | 254 Tyr> Cys | 0.8 | 0.2 | 0 | 0.9 | 0.1 |
| 5791 C>A | Intron8 | Transversion | - |  | 0.2 | 0.2 | 0.6 | 0.3 | 0.7 |
| 6303 A>G | Intron9 | Transition | - |  | 0.2 | 0.2 | 0.6 | 0.3 | 0.7 |
| 6494 A>G | Intron10 | Transition | - |  | 0.8 | 0.2 | 0 | 0.9 | 0.1 |
| 6732 A>G | Intron10 | Transition | - |  | 0.6 | 0.2 | 0.2 | 0.7 | 0.3 |
| 6908 A>G | Intron10 | Transition | - |  | 0.2 | 0.2 | 0.6 | 0.3 | 0.7 |

\* All positions respective to NCBI accession no. ON073791.1

### Association of *OLA1* mutations with heart failure

To investigate the potential association between *OLA1* mutations and heart failure, we compiled the complete *OLA1* gene sequences from each individual patient and performed alignment in MegaAlign to identify genetic variations. Subsequently, we verified each variation by cross-referencing with the respective chromatogram utilizing the Chromas Lite program. Intriguingly, we observed genetic variations in the DNA sequences of both failing and non-failing heart samples, displaying a notable correlation with failing hearts. The genotype and allele frequency patterns exhibited significant distinctions between failing and non-failing patients (**Table 1**), and these Single Nucleotide Polymorphisms (SNPs) exhibited interlinkages, as demonstrated in the MegaAlign (**Figure 3C**). The majority of these SNPs were located within intronic regions, displaying mutual correlations, implying their interdependence (**Figure 3C**).

Additionally, we identified a heterozygous exonic mutation (5144 A>G, resulting in 254 Tyr >Cys) in exon 8 (**Figure 4 A, B, C**), which has also been reported in cancer patients under the same SNP ID (rs11558990), in association with a poor prognosis. To further assess the potential impact of this mutation, we utilized various online servers for predictive analyses. The Predictor of Human Deleterious Single Nucleotide Polymorphisms (PhD-SNP) analysis indicated that this *OLA1* mutation (254 Tyr>Cys) is deleterious. Similar predictions were also obtained from PhyPhen2 (0.89) and SIFT (.078) analyses (**Figure 4D**).

Interestingly, we noted a distinct pattern of *OLA1* mutations between failing and non-failing hearts, with certain SNPs exhibiting associations with one another (**Figure 3C**). This gave rise to different haplotypes observed between the two conditions (**Table 2**). Notably, several mutations were situated in intronic regions, suggesting a potentially crucial role in RNA splicing and/or gene regulation, which warrants further confirmation.

**Table 2.** Haplotypes observed in the Human OLA1 gene.

|  | Sample | 32 | 1496 | 1573 | 2064 | 2443 | 3310 | 3792 | 4240 | 4635 | 5144 | 5791 | 6303 | 6494 | 6732 | 6908 |
| --- | --- | --- | --- | --- | --- | --- | --- | --- | --- | --- | --- | --- | --- | --- | --- | --- |
| **Non**<br>**Failing**<br>**heart**<br>**(NF)** | NF-1 | T | G | G | G | Y | A | A | G | C | A | A | A | G | G | A |
|  | NF-2 | Y | A | S | A | T | A |  | G | T | A | C | G | G | A | G |
|  | NF-3 | Y | R | G | R | T | R |  | G | T | R | C | G | R | A | G |
|  | NF-4 | Y | A | G | A | T | A |  | G | T | A | C | G | G | A | G |
|  | NF-5 | C | A | C | A | T | A |  | G | T | A | C | G | G | A | G |
| **Failing**<br>**heart**<br>**(HF)** | HF-6 | T | R | G | R | T | A |  | G | Y | A | M | R | G | R | R |
|  | HF-7 | T | R | G | R | T | A | A | G | Y | A | M | R | G | R | R |
|  | HF-8 | T | G | G | G | T | A | A | G | C | A | A | A | G | G | A |
|  | HF-9 | Y | A | G | A | T | A |  | G | T | A | C | G | G | A | G |
|  | HF-10 | Y | A | G | A | T | A |  | R | Y | R | C | G | R | A | G |

\* NF 1-5 (DNA from non-failing heart tissue) and HF 6-10 (DNA from failing heart tissue), Heterozygote code represent R-A/G, S-G/C, M-A/C, Y-C/T.

### Development of a PCR-based screening test for the *OLA1* point mutation (254 Tyr>Cys)

The non-synonymous mutation 5144A>G was identified in the *OLA1* gene in both failing and non-failing human heart patients, and it has been previously reported in a substantial cohort of cancer patients in association with poor survival. Crystallographic analysis of the mutant OLA1 protein revealed an altered structural conformation compared to the wild-type OLA1 [1]. In light of these findings, coupled with the proximity of SNP 5144 A>G (254Tyr>Cys) to the GTPase domain of the human OLA1 and highly conserved among species (Figure 4E), we devised a streamlined, cost-effective, and expeditious screening protocol for the detection of this point mutation.

To accomplish this, we developed a Tetra-ARMS PCR screening assay utilizing four distinct primer sets. The two outer primers (forward and reverse) were common for both genotypes, amplifying a 264 bp fragment. Conversely, each of the inner primer sets was allele specific. The inner forward primer designed for the G allele generated a 149 bp fragment, while the inner reverse primer tailored for the A allele produced a 171 bp fragment (**Figure 4F**). This screening protocol underwent further validation using known genotypes to ascertain its precision. As anticipated, the AA genotype exhibited two bands (264bp and 171bp), while the heterozygous A/G genotype displayed three bands (264bp, 171bp, and 149 bp) (**Figure 4F**).

Notably, this screening protocol is also applicable for detecting the *OLA1* mutation (chr2:q31.2.174082033 A>G) resulting in a 254 threonine to histidine substitution, which has been identified in cancer patients. Consequently, this screening methodology serves a dual purpose, enabling the detection of both genetic and somatic mutations, particularly the 254Tyr>His alteration in cancer patients.

To further corroborate the existence of this mutation in the human *OLA1* gene, we examined a large dataset documented in the NCBI SNPs variant dataset, along with the REGARDs study. Remarkably, the exonic non-synonymous mutation 5144 A>G (254Thr >Cys) was also observed in these datasets. This comprehensive analysis also revealed that the highest incidence of variations was located within intronic regions.

## Discussion

OLA1 is a family of GTPase proteins that possesses both GTPase and ATPase activities. Its elevated expression in cancer cells has been correlated with poor survival and clinical outcomes [1,2]. Notably, overexpression of OLA1 in cancer cells has been shown to hinder cellular apoptosis through its interaction with BRCA1 and BRCA1-associated RING domain protein (BARD1) [3–5]. Additionally, OLA1 forms a ternary complex through its interaction with elf2a, thus exerting regulatory control over protein translation and cell proliferation [6]. Recent research has revealed that OLA1’s interaction with Hsp70 is crucial for stabilizing SOD2 in pulmonary smooth muscle cells in mice. Depletion of OLA1 leads to SOD2 deficiency, resulting in increased expression of the anti-apoptotic gene, X-linked inhibitor of apoptosis (XIAP), and consequently leading to elevated proliferation of pulmonary smooth muscle cells [5,7].

In a mass spectrometric analysis of isolated proteins from failing hearts from patients with hypertrophic cardiomyopathy (HCM), the expression of OLA1 was found to be downregulated [8]. Previously, we have demonstrated that in a mouse model of angiotensin II (AngII)-induced cardiac stress, altered expression of OLA1 was shown to lead to the phosphorylation of the GSK3β/β-catenin pathway [4]. Additionally, in another independent study, whole body deletion of the *OLA1* gene in mice resulted in stunted growth, delayed development, and the birth of litters with immature organs, ultimately leading to perinatal lethality, highlighting the essential role of OLA1 in growth and survival [6]. Interestingly, this global *OLA1* knockout also led to a significant increase in heart weight relative to body weight, indicating the potential significance of OLA1 in cardiac pathophysiology [6].

In the present study, we confirmed that both mRNA and protein expression of *OLA1* were significantly downregulated in human failing heart tissue compared to non-failing tissue, underscoring the importance of OLA1 in cardiac function. Given the importance of OLA1 in heart diseases, we characterized *OLA1* at both mRNA and genomic levels and compared it with other species. Our findings demonstrated that the *OLA1* gene exhibits maximum homology with other species, suggesting its evolutionary conservation and crucial role in cardiac physiology across species. Similar observations were made for the *Tbx5* gene, which has also been shown to be evolutionarily conserved from xenopus to mammals [20].

Previous studies have emphasized the significance of mRNA splicing in post-transcriptional RNA modification, resulting in single genes encoding multiple isoforms of proteins that regulate various biological processes. Multiple isoforms of sarcomere genes have been associated with human heart failure, further highlighting the complexity of cardiac regulation [21]. Notably, the TTN gene, encoding a major sarcomere protein, produces multiple isoforms through alternative splicing, and mutations in *TTN* contributes to the development of DCM in humans [22,23]. Interestingly, the *OLA1* gene maps close to TTN at the same locus (chromosome 2, locus 1q31) associated with DCM. Our study also identified multiple mRNA splice isoforms of *OLA1* in human cardiac tissue, warranting further validation of their functions in cardiac physiology.

DCM is characterized by progressive dilation of the left ventricle and systolic dysfunction, ultimately leading to heart failure. It is the most genetically heterogeneous form of cardiomyopathy, with multiple forms of the disease linked to the same gene. This underscores the importance of genomic context in the pathophysiology of disease-associated variants. Gene mutations represent a significant cause of DCM, with over 40 genes encoding components of the sarcomere, cytoskeleton, or nuclear lamina, implicated in DCM in humans. Among these, the *TTN* gene, located on chromosome 2 at the locus 1q31, is associated with familial DCM [18]. Recent studies have demonstrated that truncation or mutation in *TTN* is a major cause of DCM, accounting for up to 25% of cases in humans [24]. Intriguingly, the *OLA1* gene is also mapped to the same locus (1q31 of chromosome 2), in close proximity to *TTN*. Furthermore, recent studies have indicated that mutations in *OLA1* are associated with poor survival and an increased risk of developing atherosclerotic diseases in humans [1,3,17]. However, it may be that the cardiac pathobiology associated with the two genes may be different. These results highlight the significant role of *OLA1* in cardiac function.

The effects of *OLA1* mutations on heart failure in human patients is yet to be studied. In this investigation, we screened for mutations in the *OLA1* gene and explored their association with human heart failure. Our findings revealed 15 mutations in the *OLA1* gene among both failing and non-failing patients. These mutations were primarily located within intronic regions, with one exception of a nonsynonymous 2144 A>G change resulting in an amino acid alteration from 254Tyr to Cys. This mutation, reported in the SNPs database under SNP ID rs11558990, exhibited a strong correlation with rapid disease progression and poor survival in cancer patients. Additionally, in the REGARDS study, individuals with the same mutation was observed within the enrolled population [25]. Our study also identified the same mutation in the *OLA1* gene among patients with heart disease. This point mutation could have a strong negative impact on heart function is in line with another point mutation in *OLA1* (E168Q) which failed to form a complex with BRCA1 and BARD1 proteins and had poor outcome in breast cancer [3]. Given the importance of the 254Tyr>Cys point mutation, we developed a rapid, reliable, and cost-effective genotyping protocol using Tetra-ARMS PCR [12,13,26]. This PCR-based assay is also useful for screening somatic point mutations in cancers as well, highlighting the application aspect of the test. Thus, overall, our findings shed light on the potential role of OLA1 in cardiac physiology and underscore the significance of *OLA1* mutations in heart failure.

## Study limitation

To establish a robust association between OLA1 mutations and heart disease, it is necessary to incorporate a larger number of human samples for further investigation.

## Data availability

The datasets used or analyzed during the current study are available from the corresponding author and Praveen Dubey upon reasonable request.

## Author contributions

PK and PKD conceptualized the initial project and study designed. PKD and SD performed the experiments and drafted the manuscript. SS, PDB, SP and PK helped revising the drafts and provided critical insights and interpretations. All authors discussed the results, interpretation, and commented on the manuscript at all stages.

## Supplemental Materials

Supplementary primer Table S1

Supplement Figures S1 – S4

## Data Availability

All data produced in the present study are available upon reasonable request to the authors.

## Supplementary Figures

**Figure S1.**
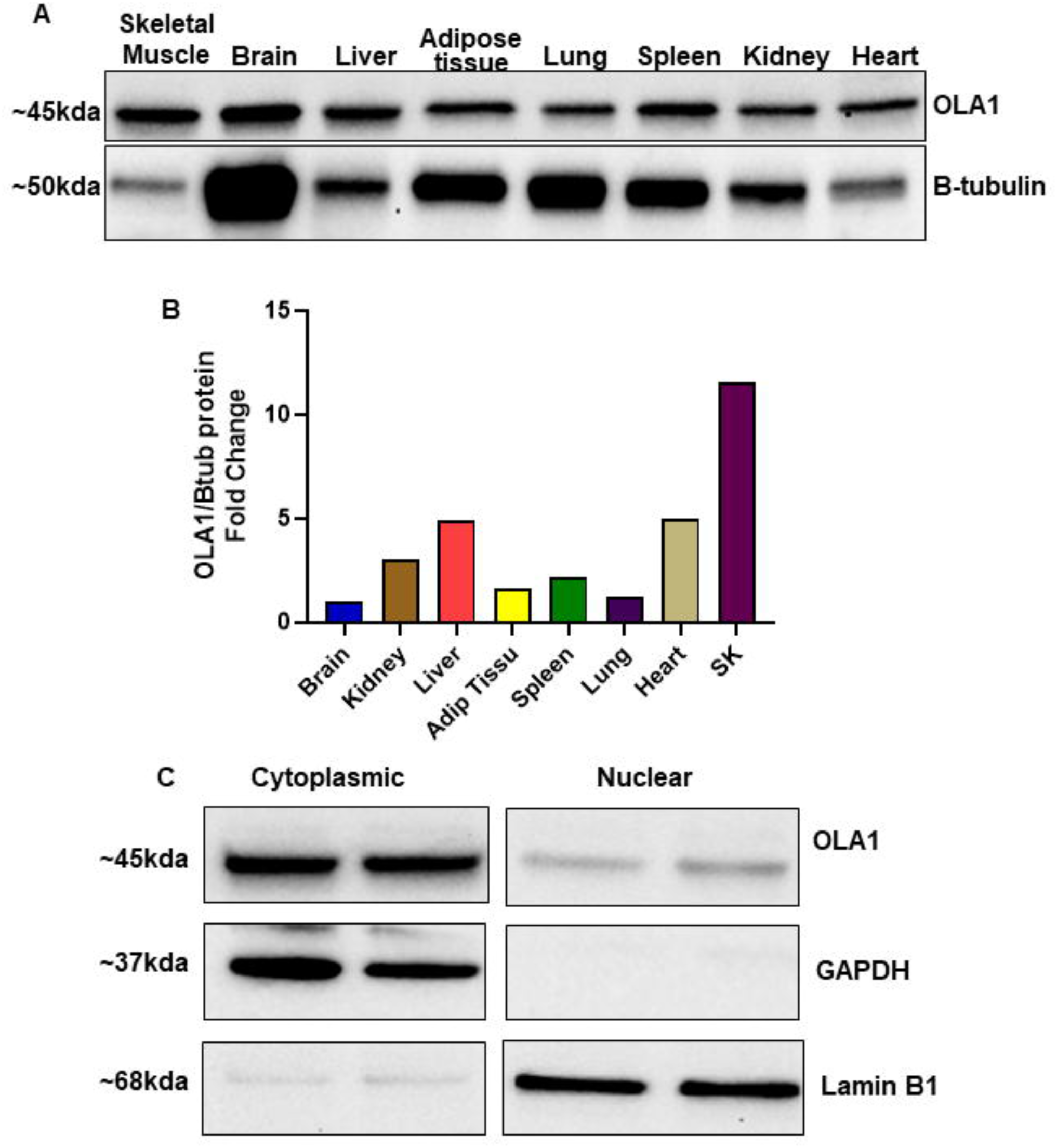
Expression and Localization of OLA1 in Mouse Tissue and Ac16 cells. (A) Immunoblot showing the expression of OLA1 and β-Tubulin in various mouse tissues. (B) Densitometric normalization of the OLA1 protein. (C) Immunoblot demonstrating the cytoplasmic enrichment of the OLA1 protein.

**Figure S2.**
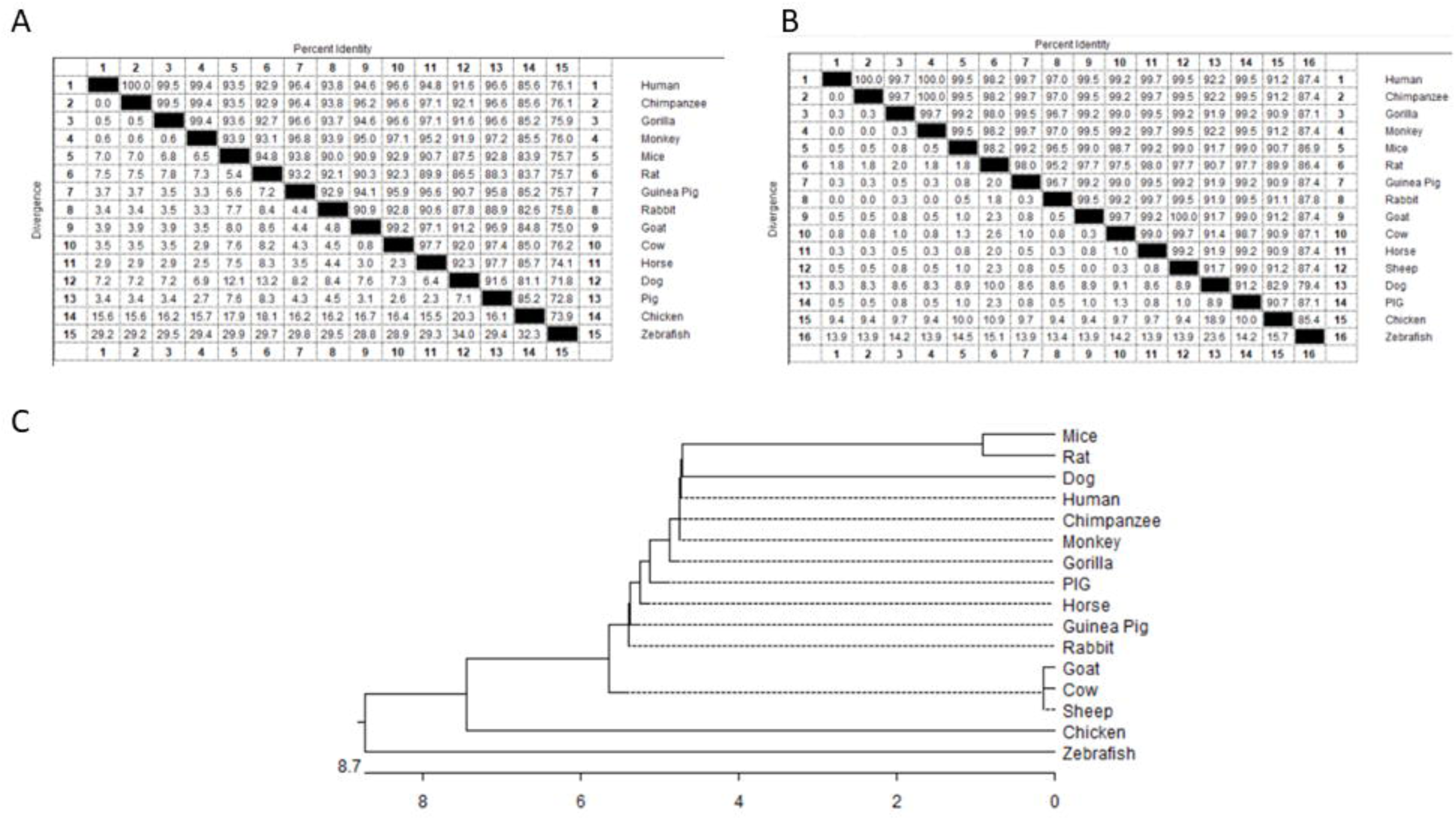
Percentage homology and phylogenetic analysis of human OLA1 gene with different species at nucleotide and amino acid basis. (A) Percentage identity of human *OLA1* with other species at nucleotide level. (B) Percentage identity of human OLA1 with other species at amino acid level. (C) Phylogenetic analysis revealed OLA1 gene are evolutionarily conserved among different species.

**Figure S3.**
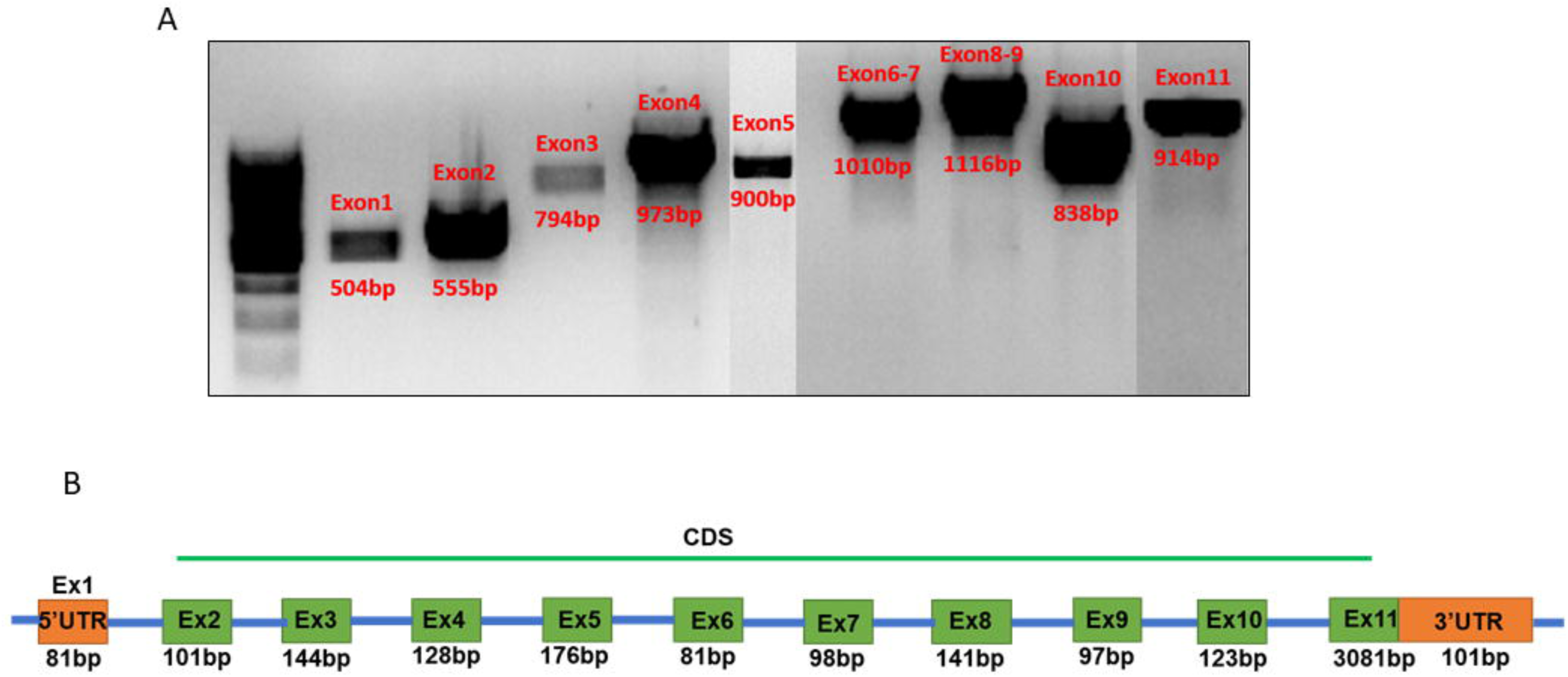
(A) PCR amplification of different fragments from the *OLA1* genomic region covering exons with adjacent introns. (B) Genomic organization of the *OLA1* gene. Ex (Exon), UTR (Untranslated Region), CDS (Coding DNA sequence).

**Figure S4.**
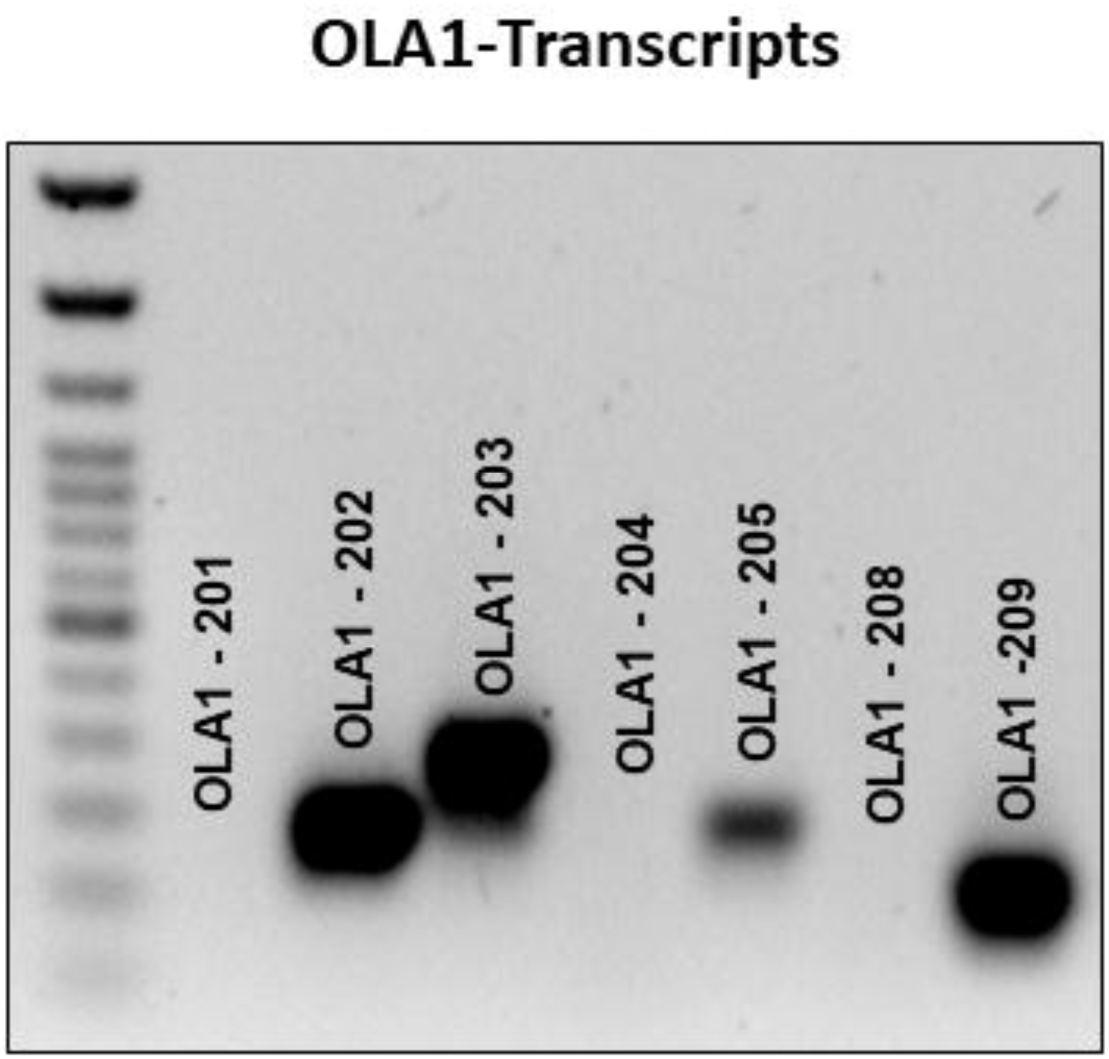
Semi quantitative Amplification of different transcript variants of the *OLA1* gene expressed in the human heart.

**Figure S5.**
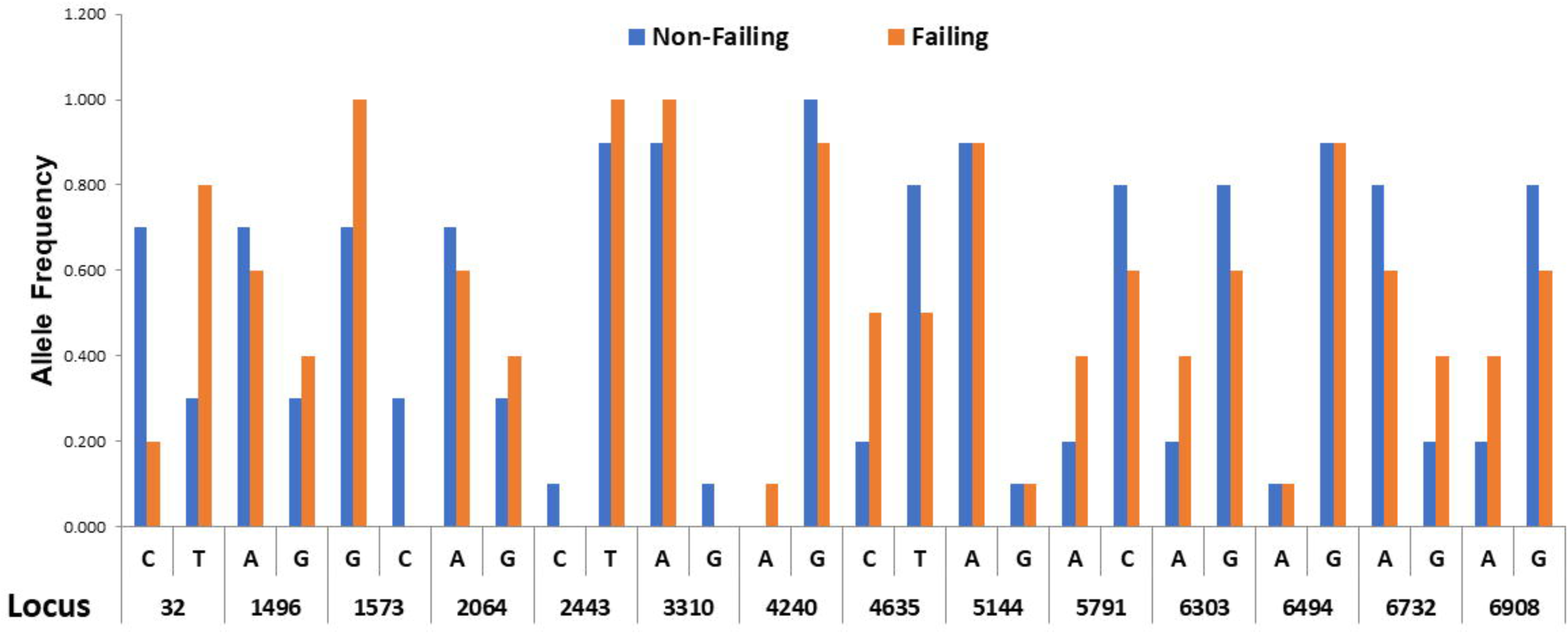
Distribution of SNPs among failing and non-failing heart patients.

**Supplementary Table S1.**
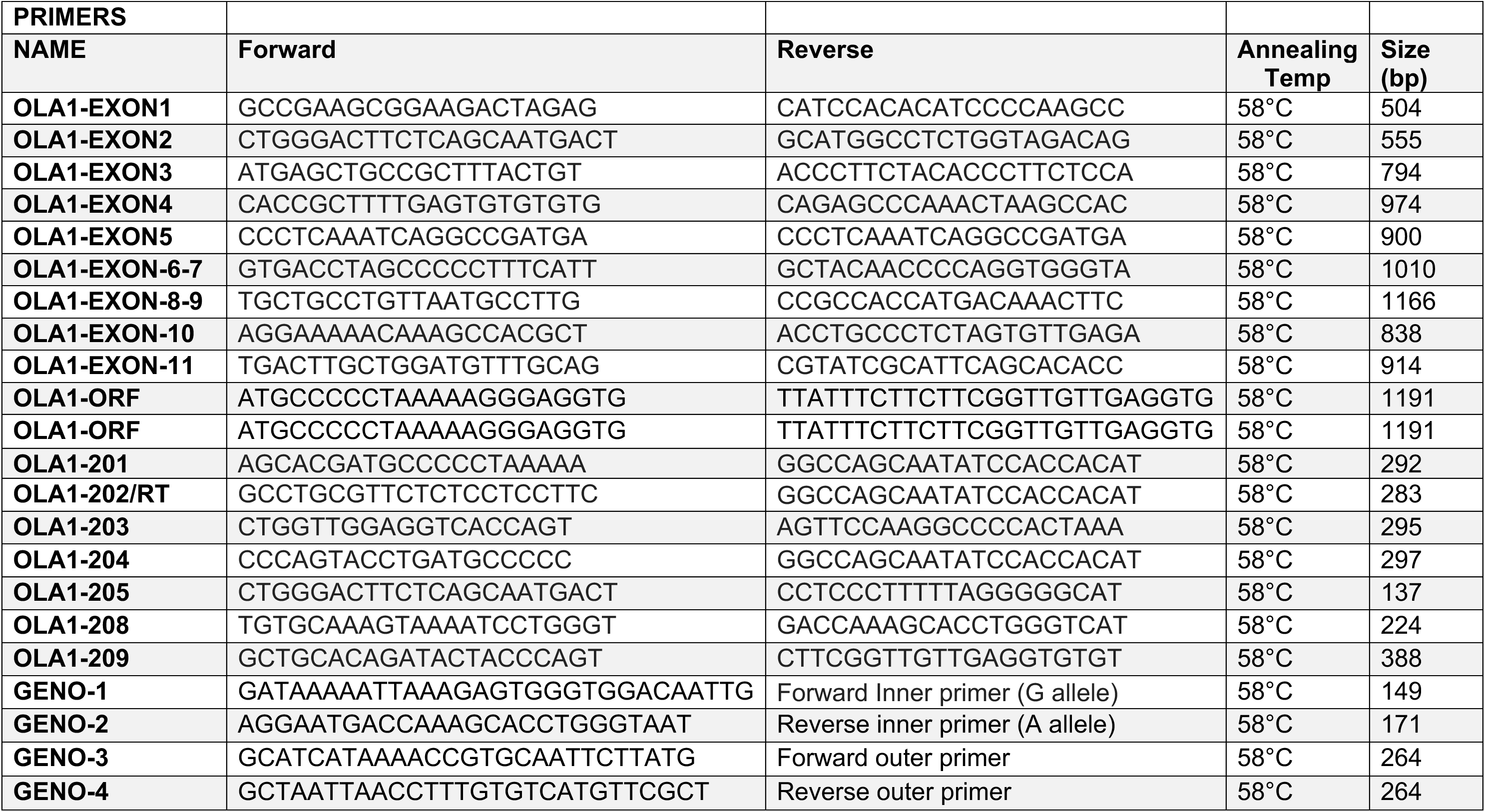
List of Primers used in the study.

